# Perceptions of health-related quality of life among heart transplant recipients: a qualitative study

**DOI:** 10.1101/2025.03.11.25322903

**Authors:** Redouane Mahmoudi, Pascal Battistella, Laurent Sebbag, Latame Komla Adoli, Francis Guillemin, Cécile Couchoud

**Author notes:** Corresponding author : Redouane Mahmoudi Agence de la biomédecine 1 avenue du Stade de France 93212 SAINT DENIS LA PLAINE CEDEX FRANCE.

## Abstract

**Background:** There is a need to improve knowledge of the health-related quality of life (HRQoL) in the post-heart transplantation (HTx) period and the factors affecting it. This qualitative study aimed to identify the most important domains of HRQoL for heart transplant recipients and the factors that impact it.

**Methods:** This was a qualitative study across 5 geographically diverse large HTx centers in France from July 2022 to January 2023. We gathered a purposive sample of individuals who had undergone HTx. A face-to-face semi-structured interview guide was used for individual interviews. All interviews were audio-recorded and transcribed verbatim.

**Results:** A total of 14 individuals (10 men) were interviewed. The data analysis led to the development of 8 main themes (with sub-themes) that were relevant to participants: HRQoL perception (mental health, physical capacity, symptoms and comorbidities), participants’ experience during the HTx process, immunosuppressive treatments, relationship with the healthcare team, external and internal resources, socio-economic aspects and feelings about the donor. Recipients, spontaneously made connections between these themes.

**Conclusions:** Heart transplant recipients had diverse perceptions of their HRQoL in the post-HTx period. The rich variety of themes identified from the review highlights that recipients have a complex HRQoL profile which is not currently captured by standard HRQoL tools that are commonly employed. These aspects should be taken into account in the clinical follow-up and in the selection of the most appropriate Patient Reported Outcome Measures (PROMs).

## INTRODUCTION

Heart transplantation (HTx) remains the best therapy for terminal and refractory heart failure (1,2). It increases the life expectancy and generally improves patients’ health-related quality of life (HRQoL)(3).

This improved HRQoL is stable over time (4) and can be seen in different areas of quality of life (QoL) such as physical or mental aspects but also social and family aspects (5). However, HRQoL can be compromised by specific complications such as heart rejections, infection, graft failure, cancer and limited survival rates, which may affect HRQoL (6).

In all cases, HTx recipients seem to have lower HRQoL than the general population (7), because HTx affects recipients’ activities of daily living and presents various challenges such as those related to their new lifestyle (8,9). Considering these aspects in the clinical follow-up or in the evaluation of HTx benefits seems fundamental. It helps promote patient-centered care. Assessment of HRQoL is also developing into a new medical indicator in transplantation medicine (10).

HRQoL is defined as the aspects of quality of life that are directly or indirectly related to health. It encompasses a broad range of concepts, including disease symptoms of disease, health conditions, treatment side effects, and functional status in physical, mental heath, and social domains (11,12). For HTx recipients, HRQoL also reflects the impact of transplantation and immunosuppressive treatment on their level of disability and daily functioning.

There are various generic and condition- or transplant-specific patient-reported outcome measures (PROMs) available to assess HRQoL. But, there is no gold standard instrument that meets all recipients’ expectations and that is easy to use, reliable and valid (13). They may focus on one or more aspects of life: overall HRQoL or specific domains, such as physical capacity, psychological distress, ability to participate in social roles and activities and treatment side effects.

The French Agency of Biomedicine wishes to organize the monitoring of the HRQoL of transplant recipients as part of its national transplant registry in order to identify actions to be implemented. Individual semi-structured qualitative interviews enable a deep understanding of the patient’ experiences and provide useful information about their perceptions, beliefs, and attitudes and how they view their HRQoL (14). We aim to conduct an in-depth exploration of HRQoL among HTx recipients. Through individual interviews, we seek to identify key themes that impact their HRQoL, including physical, psychological, and social factors. This qualitative data will help us gain a deeper understanding of the experiences of these patients, enabling us to select or refine existing questionnaires for a more accurate measurement of HRQoL in this specific population and to tailor care accordingly.

## MATERIALS and METHODS

### Study design

This was a descriptive cross-sectional semi-structured qualitative interviews .

### Participant recruitment

Patients were recruited in 5 university hospitals in France. The inclusion criteria were 1) isolated HTx recipient; 2) age ≥ 18 years and scheduled for a routine outpatient consultation; 3) able to read, write, and speak French; 4) physically able to participate in the study; 5) able to give informed consent ; and 6) willing to participate to the study.

Staff members referred patients who met the study criteria to a research team member to discuss the study in detail and confirm their willingness to participate in the interviews. Each patient who came to consult on a given day was given the choice to participate or not. All included patients agreed to have individual interviews. They were informed that they could withdraw from the study at any time until publication of the findings.

### Data collection and interview process

The themes of the interview guide were determined by the results of our previous literature review (13). Therefore, our guide covers the following topics: current and future health status, patient caregiver relationship, health-related quality of life, and immunosuppressive treatment **(Table 1)**. The questions on these topics in the interviews were open-ended. The semi-structured format allowed recipients to tell their experience in the manner they chose (15). Therefore, the guide questions were not necessarily asked in sequential order.

**Table 1:**
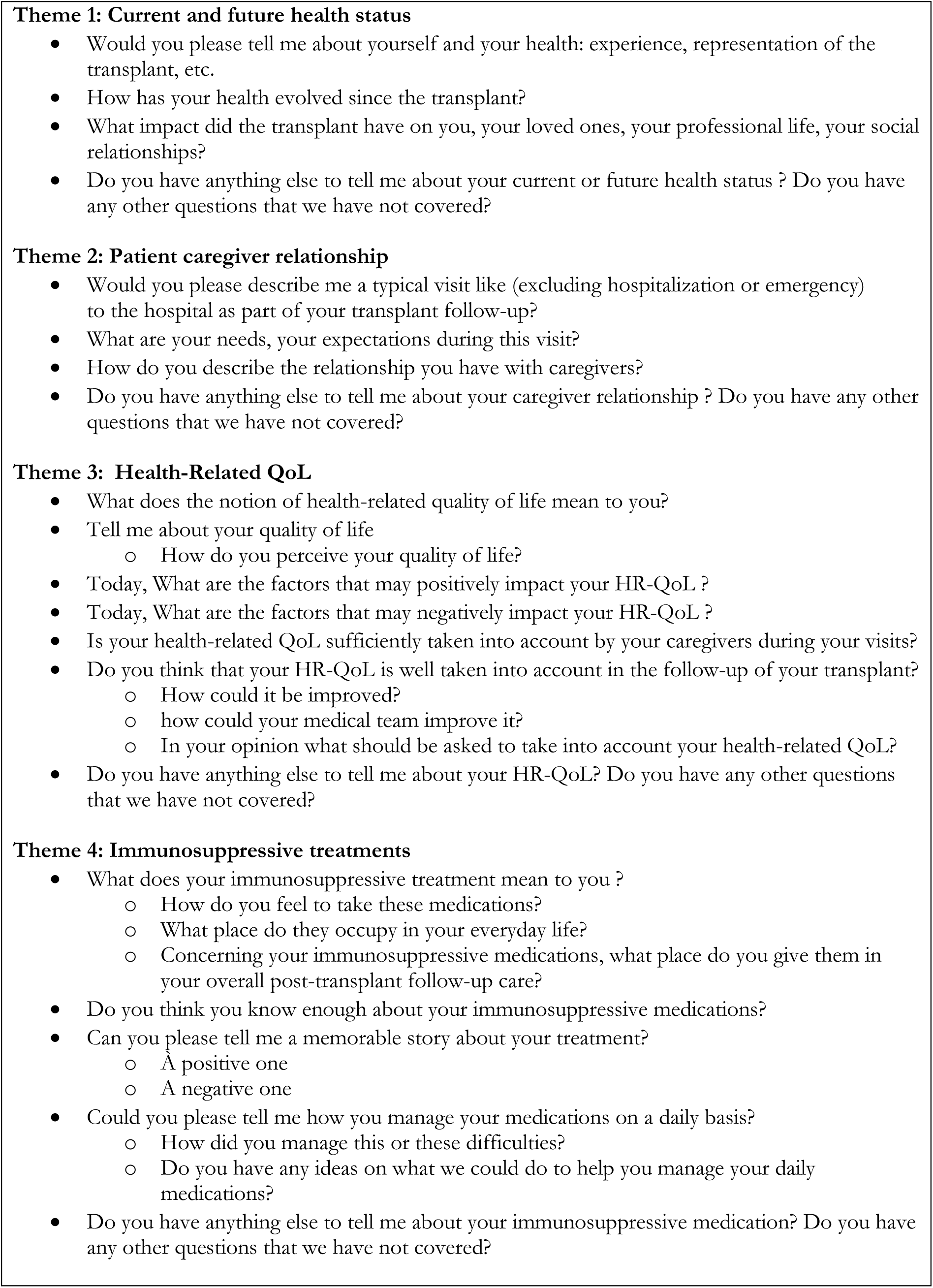
Semi-structured interview guide

The interviews took place in an isolated location in the hospital (office, meeting room) to maintain participants’ privacy and data protection (16). Patients were asked about their general health, their HRQoL, how they cared for their medical conditions, and the impact of immunosuppressive treatment on their lives.

Data were collected between July 2022 and January 2023. The interviews were conducted individually, in French, and took place at the hospital where clinical follow-up was performed. The interviews were conducted by RM, who is qualified to do so.

A few minutes was spent before the start of the interview to establish rapport and trust with the participant and ensure confidentiality. All interviews were recorded and transcribed verbatim (including dialog, pauses, silences and laughter) by using Microsoft Word software after each interview. Notes and observations of the interviewer were kept to capture all clues from the non-verbal communication.

### Ethical considerations

The participants were informed that the interviewer was not a member of the healthcare team. Participants were aware that the interviews were transcribed and anonymized and that none of the clinical staff would have access to the transcripts. They were informed that they could ask to stop the audiotape recording during the interview or that they could refuse transcription of some parts of the interviews. Furthermore, all participants signed an informed consent form before the interview.

In accordance with French law, research studies based on the national organ registry (CRISTAL), managed by the Agence de la Biomédecine, are considered part of transplant assessment and do not require additional Institutional Review Board approval. The database has been registered with the French National Commission on Computing and Liberties (CNIL).

### Data analysis

The Consolidated Criteria for Reporting Qualitative Research (COREQ) checklist was used as a reporting guideline for this study (17). Data were analyzed using inductive thematic analysis to identify themes of related HRQoL post-HTx (18). The analysis was conducted iteratively, with researchers identifying units of meaning, coding all meaningful text into concepts derived inductively from the data. Similar concepts were grouped into preliminary sub-themes and broader themes, and any unclear codes or themes were discussed until consensus was reached (15). Concept saturation was documented using a saturation grid. To enhance investigator triangulation each written transcript was analyzed independently by RM and CC (female, MD, PhD, have knowledge in qualitative studies, not involved in patient care). All analyses were conducted without the use of qualitative software programs.

Themes related to the perception of HRQoL and factors influencing it, were identified. Connections between these themes, as perceived by the patients, and their literal meaning were described, including their direction and meaning.

## RESULTS

We included 14 individuals with isolated HTx, 10 men, 1 to 4 patients per facility. The mean age of participants was 62 years (range 41–81). The mean number of years post-transplantation was 14.2 (range 2.5–31). The mean interview time was 51 min (range 34–80) (**Table 2**). After the 12^th^ interview, there were no new themes generated from the interviews. We conducted two additional interviews to ensure and confirm that no new themes were emerging (19,20).The transcription of these 14 interviews identified 8 major themes mentioned by the recipients. External and internal resources took a big place in our interviews because patients constantly addressed this theme during various questions that did not specifically relate to it. All themes and sub-themes are further described and illustrated using patient quotes from the interviews (**Table 3**). Based on patients’ spontaneous expressions, we identified links between the HRQoL theme and seven other themes that are likely factors influencing HRQoL. These relationships can have a dual effect, either positively or negatively impacting HRQoL (**Figure 1**). The connections between the themes emerged directly from the associations spontaneously expressed by the patients.

**Table 2:** Patient demographics

| **Participants** | **Sex** | **Years of heart**<br>**transplantation**<br>**(Y)** | **Length of**<br>**interview (min)** | **Time since**<br>**heart**<br>**transplantation**<br>**(Y)** |
| --- | --- | --- | --- | --- |
| Patient 1 | F | 2004 | 68 | 18 |
| Patient 2 | F | 2020 | 45 | 2.5 |
| Patient 3 | M | 1991 | 37 | 31 |
| Patient 4 | M | 2014 | 48 | 7.8 |
| Patient 5 | F | 2015 | 34 | 6.8 |
| Patient 6 | M | 2012 | 44 | 10.4 |
| Patient 7 | M | 2000 | 34 | 21.6 |
| Patient 8 | M | 2019 | 34 | 3.2 |
| Patient 9 | M | 1999 | 59 | 23.3 |
| Patient 10 | M | 2000 | 56 | 22.4 |
| Patient 11 | M | 2016 | 80 | 6 |
| Patient 12 | M | 1997 | 62 | 24.7 |
| Patient 13 | F | 2015 | 35 | 6.6 |
| Patient 14 | M | 2008 | 59 | 15 |

**Table 3:**
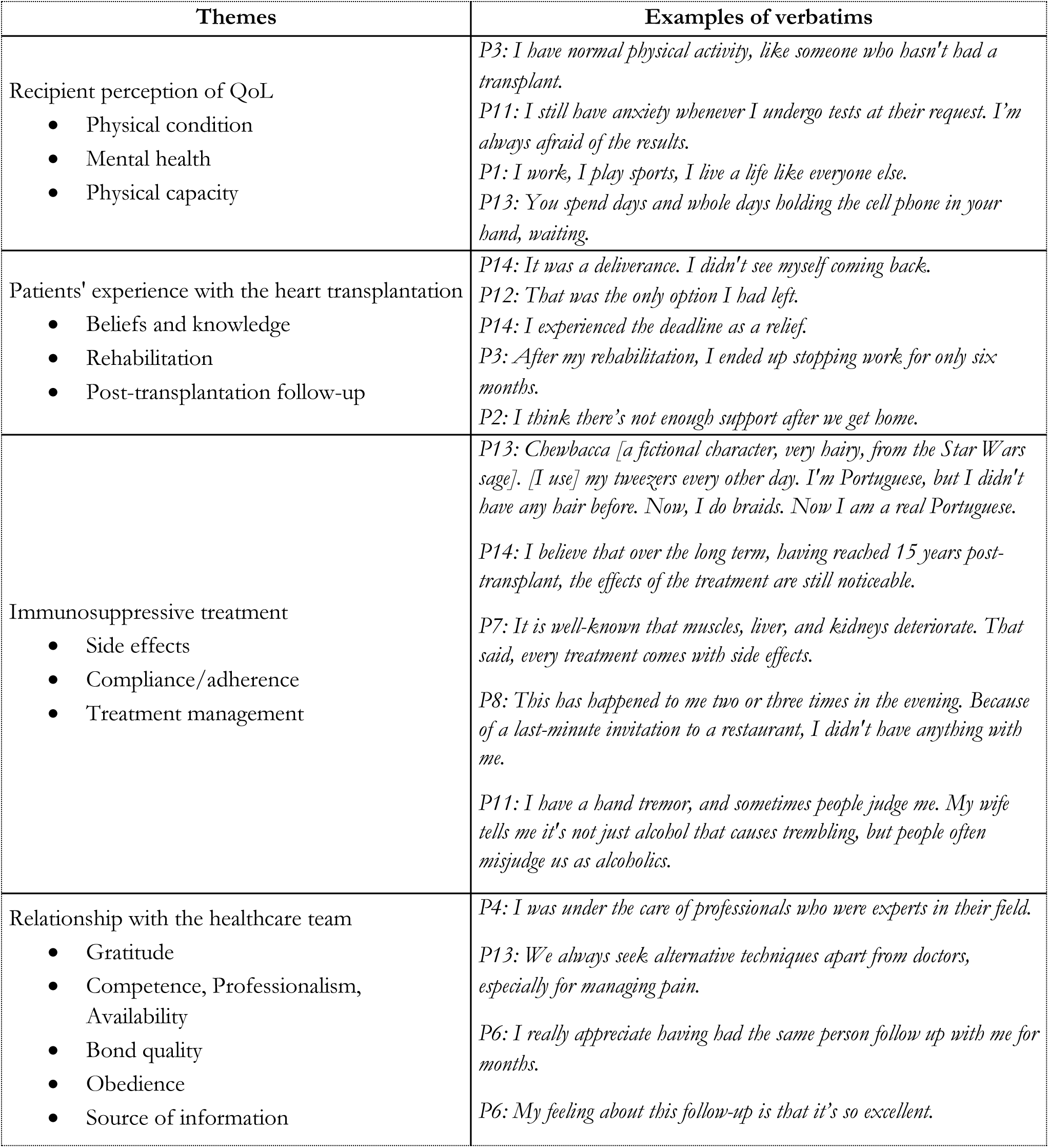
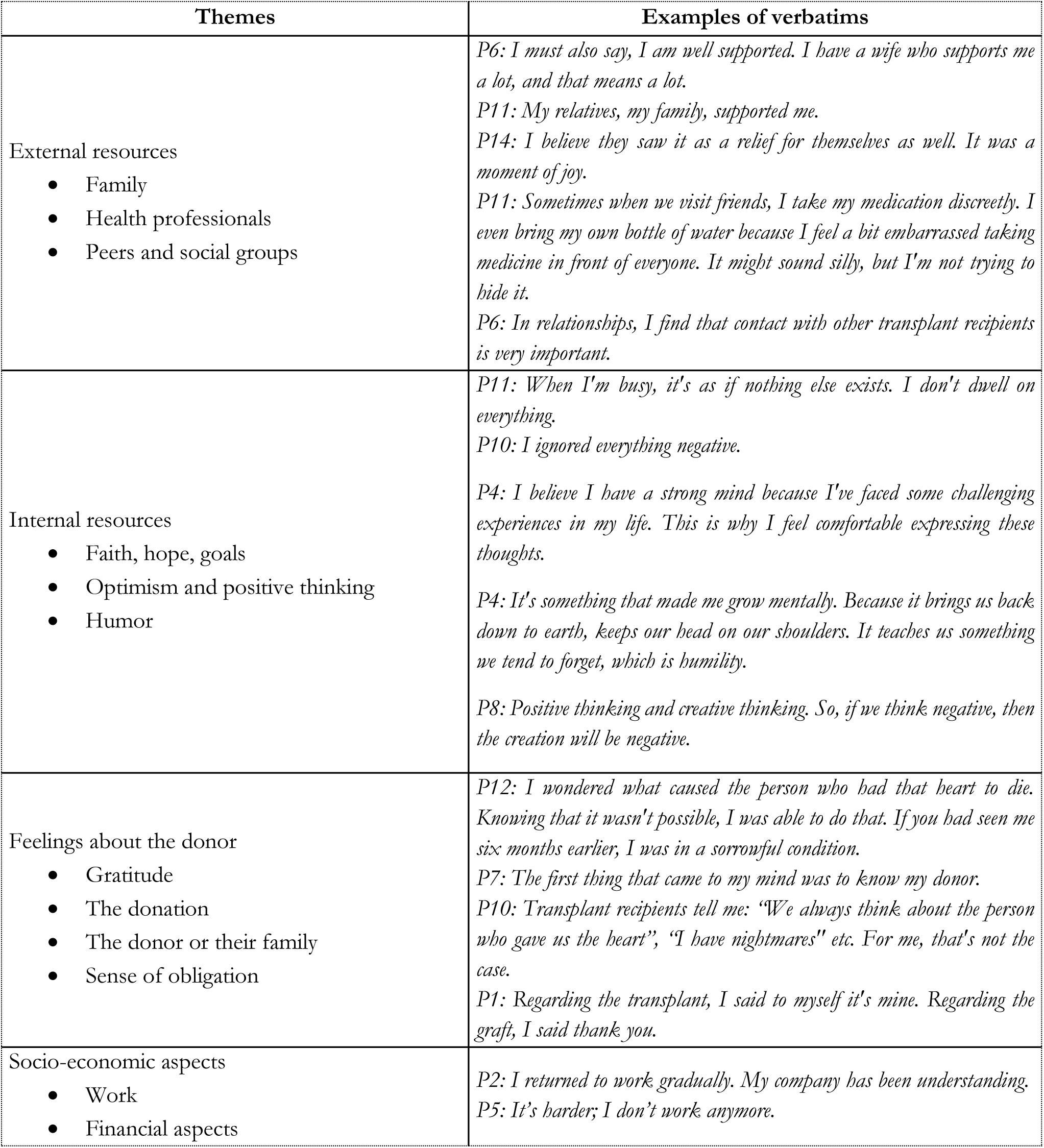
Themes, subthemes, a selection of illustrative verbatim quotes

**Figure 1:**
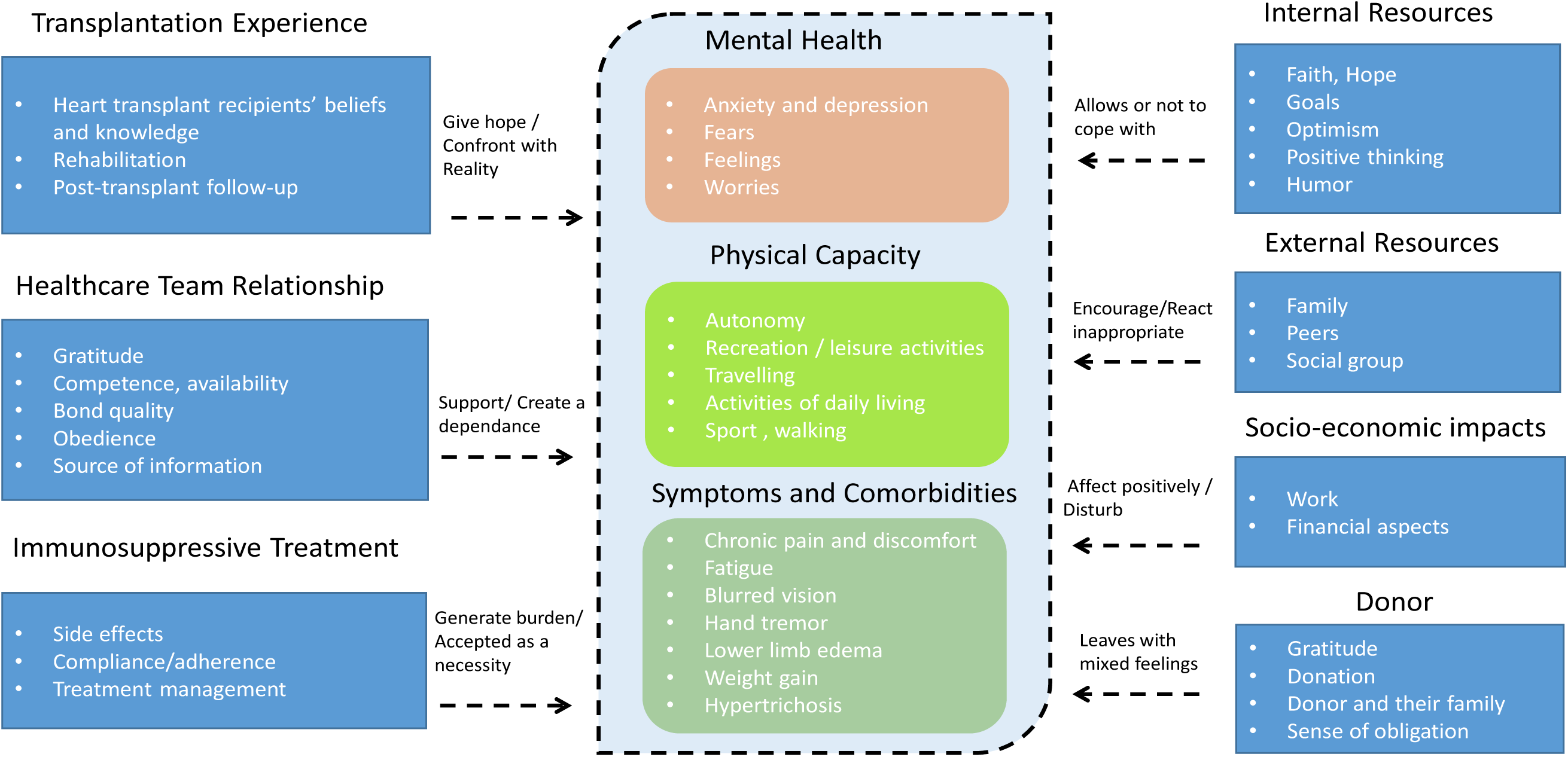
Mapping of HRQoL

### Components of health-related quality of life

Three subthemes emerged from the data, constituting a broader category of recipients’ HRQoL perception. Recipients reported significant impacts on their mental health, physical capacity, and symptoms when compared to their life before HTx and/or before their heart disease onset. While these three areas represent only a facet of HRQoL, they were identified as the most crucial aspects for patients. Additional domains have also contributed, both positively and negatively, to the overall HRQoL. All domains are interconnected and may influence each others (figure) .

#### Mental health

Undergoing HTx is an emotional experience and some patients found it very stressful and felt anxious and depressed. For some patients, this stress remained even after the HTx, especially when medical tests (endocardial biopsy, blood analyses, etc.) are required and may have negative results. Experiencing uncertainty and mental health problems negatively affected their HRQoL. *“P5: I am always anxious and stressed”,* “*P1: I am always afraid. It’s a source of stress*”.

#### Physical capacity

This refers to the ability to perform daily activities such as walking, driving, working or leisure activity. The search for autonomy takes a center place. Limitations in activities and social interactions resulted in negative feelings. However, many participants highlighted a restored ability to participate in activities of daily living, such as gardening, doing odd jobs, traveling or practicing a sport. This restoration was thought to have a positive effect on HRQoL. *“P2: Go shopping, go to a store alone, buy clothes. Before, I always had to be accompanied by my children. Now I am becoming independent again”,“P3: The key aspect is to return to a normal life, which includes working, family, and maintaining a social life”*.

#### Symptoms and comorbidities

Many symptoms are linked either to the initial illness, side effects of the immunosuppressive treatments or aging. Recipients mainly experienced fatigue, muscle and joint pain. *“P14: Efforts, intensive efforts, long effort. I can’t handle heavy tasks”, “P14: Limiting is the effort, fatigue during exercise and muscle and joint pain.”*

### Patient’s experience with the HTx

Patients identified the pre-transplant period as a journey toward certain death. Therefore, undergoing HTx was described as “*an extraordinary thing*”, “*a renaissance*”, “the only way possible”. *“P3: I knew that it was the only option available”,”P10: It’s a rebirth. If I hadn’t had this, I wouldn’t be here anymore”*.

Most patients describe the transplantation as a beautiful experience. They were satisfied with it and had positive perspectives toward it, although they experienced a longer recovery process than expected, punctuated by stress, episodes of rejection, and other diseases linked to the HTx or immunosuppressive treatment. In general, recipients felt better than expected but worse than before their cardiac illness. *“P2: I think about the transplant when I see my scar and when I take my medication differently the rest of the day…no”. “P4: I’m good, I’m really good, in my life I’m really good and I’m alive”*.

These feelings were often associated with gratitude toward the donor and his family or toward the healthcare team. All recipients immediately accepted the graft as their own. Only one patient avoided talking about it and thinking about the transplant and the donor and expressed regret for consenting to HTx because of the resulting suffering, although such feelings were balanced by the satisfaction that life had been extended. *“P13: Honestly, if I had been aware of all that, I wouldn’t have agreed to it. Considering everything that’s happening, all the health issues it’s causing… If I had known, I wouldn’t have undergone this transplant”*.

### The burden of Immunosuppressive treatment

Participants highlighted the burden of immunosuppressive treatment. Issues regarding medication management and medication-associated adverse effects emerged as major barriers to good HRQoL. Despite being informed by the healthcare team of the side effects of such treatment, some patients were not prepared and were unpleasantly surprised by the side effects. Side effects such as weight gain, hand tremors, memory lapses, and hirsutism have deleterious effects on patients’ body image and emotional functioning. Such side effects often caused them to feel anxious and uncertain about the impacts of medication on their body. Long-term complications such as skin cancer, kidney and liver dysfunction remained a source of concern. Only one patient reported that immunosuppressive treatments did not cause any side effects or discomfort*. “P14: The first concern is rejection and the anti-rejection treatments. Following that, there are treatment effects, such as increased vulnerability to infections.”, “P13: It can affect the kidneys, it can affect the liver, it can affect the bones, all that. I think there is a lack of communication on this.”, “P7: But what they never mention are the side effects, which are very difficult to tolerate”*.

Some patients reported having difficulty in organizing medication intake (preparation of the pill box and supply of drugs), whereas others saw it as a necessary evil and took their treatment like any other therapy “P2 *: Sometimes it can be restrictive. For example, I might say Saturday and Sunday when the alarm rings at 8 a.m. and you have to get up even though you’re sleeping*.”, “P13*: When we go on vacation or on the weekend, it requires quite an organization. It’s chaotic*.”, “*P6: In other words, I am very diligent about taking it almost exactly on time, but I approach it like taking a Doliprane (i.e., acetaminophen). I don’t let it define an illness for me*”.

Patients were motivated to take medications to maximize graft survival and described them as “*indispensable to life*.” *“P2: Well, if there is no treatment, there is no life”, “P4: My treatment keeps me alive. Without it, ciao bambino”*.

Most patients claimed to be compliant with their immunosuppressive treatment, and as proof, they considered taking immunosuppressive treatment as “a *habit*”, a natural “reflex”. *“P7: It’s a ritual. It’s logical. It’s just routine.”, “P8: It’s like eating, drinking, or something similar”*.

While recipients were aware of the importance of compliance with immunosuppressive treatment reality was more complicated than it seemed. *“P4: If I ever forget my pill box, I immediately turn back, no matter where I am. I go home to get it. Overall, when I take [the medication], I feel satisfied.”,”P14: The treatment is restrictive. The day we find a way to undergo transplantation, to have a pump without relying on medication, it will be ideal.“*

Some patients were aware of their transplant status mainly through taking medication and little by medical follow-up and further medical examination. *“P2: I’m aware I have a new heart, and I know it’s a good match. The transplant only crosses my mind when I see my scar or when I take my medication during the day. Otherwise, it doesn’t”*.

In summary, immunosuppressive treatments induced side effects that played an important role in HRQoL and the perceived health status of HTx recipients.

### Relationship with the healthcare team

All participants described the healthcare team as efficient, professional, attentive and like a second family. *“P1: They’re like my family, my second home. They’re important to me, just like my family”,“P14: Trust, expertise. There’s also flexibility and accessibility”*.

The healthcare team was not the only source of information, because some recipients resorted to other sources of information such as searches on the Internet, reading medication instructions or seeking information from other recipients via the Internet or discussion groups, especially regarding the side effects of immunosuppressive treatments. *“P4: I seek additional information like everyone else, using the Internet if I need something”, “P10: I don’t need to go elsewhere for information. I have access to all the specialists here”*.

Patients expressed profound gratitude to the commitment to their aftercare by caregivers. There was also gratitude to the HTx team. *“P1: I thank them again and again. I don’t know how many times I can thank them.”*

Many patients shared their point of view regarding trust and compliance with the healthcare team. *“P4: My job was to go home. And to get home, we had to listen to what we were instructed to do. You have to listen to the doctors.”. “P9: I tell you; you have to listen to them and apply the advice they give you.”*

A good relationship with the healthcare team contributed to a better perception of HRQoL, and patients were more likely to comply with their treatment with a good relationship, including complying with healthcare team instructions.

### External support (from close circle, peers and social groups)

External support and social interactions were an important theme with regards to HRQoL

#### Support from close circle

Recipient support persons were more likely to be the patient’s spouse, partner or child. There was no report of support from friends or work colleagues. Some patients experienced negative aspects because they felt like they were a burden to others, especially their family. *“P6: My children and my wife did not understand. They told me: “you had a transplant and everything and you don’t look well”*.

However, most had good relationships or connections and considered them important for HRQoL. *“P2: My daughter took my place for a year, cooking with my husband, doing housework too, with her brother and everything”,”P9: I had significant support from my family… So, this positive family aspect has been gradually strengthening since my transplant”*.

Patients described those around them, mainly their spouse, partner and children, as extremely overprotective. Relatives were very worried and did not always understand them. *“P5: Yes, my husband … prepares [the medication] for me. (Laugh). He’s so scared I won’t take [it] that he’s the one who prepares everything for me.”. “P3: Initially, they were more focused on slowing me down”*.

Recipients’ loved ones considered them fragile because of their HTx and/or the immunosuppressive treatment. *“P11: My wife was afraid. She was more aware than me that a microbe, an infection… We are fragile.”. “P2: I am quite protected, monitored. I have an individual office since the occupational doctor doesn’t want me to be with everyone.”*

Patients considered themselves, for the most part, as fragile and more likely to get sick. *“P1: I’m not immune, so it’s complicated.”. “P4: The smallest thing can be extremely, extremely serious for me.”*

The interviews highlighted a high level of stress and anxiety among recipients’ loved ones, before and after the transplant. *“P2: During the first months, after the transplant, yes, they were quite worried.”. “P3: I think it’s more difficult for those close to me than for me.”*

The participants experienced the staring and curiosity of others, from family, friends and colleagues. Some mentioned that how others viewed them had changed after the HTx, which can cause frustration. They suffered the incomprehension of relatives, the curiosity of outsiders and the stares of work colleagues*. “P2… people came to ask me lots of questions. So sometimes it makes me happy, and other times I feel like I’m seen as a curiosity”, “P3: In certain workplaces, disclosing that I’ve undergone a transplantation can be difficult and can also affect my career growth”*.

#### Support from peers and social groups

Some recipients mentioned the need to talk to peers during physical exchange groups or on the Internet. They shared their experiences particularly during rejection episodes and about immunosuppressive treatment. They felt supported and belonging to a community. Despite the curiosity, sometimes unhealthy, and the fears that this curiosity generated, the participants maintained these links through dialogue and information. *“P6: It’s important not to feel alone as a transplant recipient, but to connect with others who have had the same experience”,* “*P13: If I want more information, I use a Facebook group where we share tips and talk to each other*”.

Finally, maintaining a strong familial relation and social participation were considered important for a positive HRQoL. Also, the support from health professional was important and implied their availability.

### Internal resources

Many recipients used internal resources to help them cope with their situation. Mobilisation of internal resources included using humor, looking for information about medications, trying to distract oneself with leisure activities, thinking positively, thinking of good things, trying to keep life normal, focusing on faith, or in contrast, accepting the situation or being resigned (21). *“P4: I manage to make my mother laugh in my hospital room”. “P14: I follow medical news”. “P1: You still have to be positive in these cases, despite it being all negative”. “P11: Sometimes I go out into nature, ride my bike and then take a break”*.

Recipients noted that they tried to live in the present moment meaning no longer worrying about what happened in the past (health status before the transplantation) and not fearing what will happen in the future (side effects, rejection). *“P12: For me, reality matters a lot”, “P9: Life is about the past, but it’s mostly about the future.“*

HTx results in major stressors such as anxiety, depression, and fear. Recipients tried to maintaining hope and keep the faith.*“P4: We need something to hold on to. There are times when we face serious health concerns. We need something to hold on to”,“P14: There’s spirituality and philosophy, which can be considered”.* Recipients used acceptance or resignation to deal with the difficulties linked to post-HTx follow-up or the side effects of immunosuppressive treatments. *“P10: I suffered my case; I suffered my case”,“P8: You have to be in acceptance”*.

Personality traits, particularly a strong character or positive personality traits were highlighted as helping heart recipients to deal with adversity. *“P1: Fortunately, I have a strong character. Without it, many people might have given up”,*“*P14: Character still matters, because we experience phases and periods that can be quite challenging”*.

One patient raised the importance of making peace with oneself and others to move forward. *“P4: The fight of our life is the fight we wage against ourselves. And there I achieved peace with myself. I succeeded. But I managed to make peace with myself”*.

One recipient reported that his transplantation would benefit future transplant recipients. *“P12: I say that what they have done to me will be useful for those who come after. I am here to advance science”*.

Despite the difficulties encountered, recipients were determined to remain positive. They generally coped well and were effectively using some combination of coping strategies. Those strategies were mobilized especially for dealing with fear of the future, rejection and anxiety about accumulating side effects*. “P2: From time to time, I tell myself how long it will work. It’s more about the long term”,“P11: I still have this disease. It may be stable … it’s the future that worries me”*.

### Feelings about the donor

Heart recipients said they viewed their transplant as a gift. They were very grateful to the donor and the donor’s family. Only few viewed this gift with regret or guilt. This gift required them to do everything possible to preserve graft function as long as possible; being committed to adherence to healthcare team recommendations in particular.

The awareness of having another person’s heart inside them faded over time. *“P6: Some perceive grafts as something foreign or not belonging to them, whereas I have never experienced that feeling, not even once”*.

Many wondered about the age and sex of the donor. *“P11: I must say I always think about it”,”P5: I would like to know if it was a woman, if it was a man, if it was a young person”*.

Participants had strong feelings of gratitude for the donor and the donor’s family. *“P1: I thank these people because it was unfortunate for them, within their family. As for me, we were happy because I am here. But what they did is wonderful”.* Sometimes gratitude toward the donor was evoked with guilt that the donor died so that they could have this organ. *“P5: Well, for me, it hurts because, initially, I told myself that someone died to save my life.”*

Two patients reported frequently shedding tears when they thought of their donor. *“P11: It makes me sad; I think of him [the patient is crying]”,“ P1: He made a beautiful gesture. Tears come to my eyes [the patient is crying]. It affects me”*.

In addition to a feeling of gratitude toward the donor, recipients also had a sense of obligation (sense of duty), which consisted of taking care of themselves and preserving the graft as best as possible. *“P2: I cannot afford, in relation to the donor, to deviate”.* The heart is associated with life and often considered a source of love, emotions. *“P2: I was not at all ready for the transplantation. Because for me, since the heart is the centerpiece.”*

One recipient reported questions from those around him about his ability to love and felt resentment following his transplant. “*P12: People came to ask me questions. Since I have someone else’s heart, do I still have the same feelings? Can I still love?”*.

Contact with the donor’s family was only through the transplant coordinator. It was common to write a letter of thanks, but this was sometimes seen as very difficult. Some failed to write or just to talk about it. *“P1:I would like to write but I don’t know. I’m staring at my paper. It’s complicated. But after 16 years, I won’t do it now”,”P5:I only know I received a heart. I don’t know where it came from. It deeply touched me. I can’t talk about it”*.

### Socio-economic considerations

The financial impacts of transplantation contributed to recipients’ post-transplant anxiety and distress. Maintaining a full-time job was complicated. Some had a workplace adjustment or took early retirement or never returned to work. *“P2: I requested a change of job position. I am quite protected and monitored. I have an individual office since the occupational doctor doesn’t want me to be with everyone.”,“P14: Obviously, I wish I could have returned to my job and been able to work straight away”*.

The care of transplant patients is fully covered by the French national health insurance, but there remains a financial impact linked to some out-of-pocket costs (long-distance transport). One patient mentioned that banks refused credit or a loan because of no insurance coverage.*”P1: “I applied for a loan; it’s complicated for us too. They don’t lend to us because they don’t see us there anymore”*.

### Mapping of HRQoL

We identified factors influencing HRQoL based on patients’ spontaneous expressions, which can either positively or negatively impact HRQoL (**Figure 1**). HTx is a profound life change for patients and their families. It gives hope after often a lifetime of exhaustion and illness. Each heart recipient had a unique experience of life with HTx. The confrontation with reality is more or less complicated given the post-operative period, complications, and the recovery duration. For example, going on holiday, a strong expectation, can be problematic both due to logistical problem with the immunosuppressive treatments and global fatigue*. “P1: The transplantation allowed me to do physical activities again. I was less tired.”. “P14: The first two years were very difficult.”. “P3: finally, at the end of my rehabilitation [post-transplant cardiac rehabilitation], I stopped working for only six months”*.

Although families can be extremely overprotective, it is unfailing qualitative support with their encouragement. Certain inappropriate reactions from others could have a negative impact on their mental health. To a lesser extent, an external qualitative support contributes to the ability of recipients to participate in social roles and activities and therefore have an impact on their physical condition. *“P4: I’m really lucky to have had everyone with me”,“P6: I am well surrounded. I have a very supportive wife and that means a lot.”. “P11: My relatives, my family helped me”*.

The obedience to and the assumed dependence on the healthcare team was an important element for transplant recipients. Heart recipients expressed the fact that their HRQoL was influenced by the healthcare team’s availability and the information they gave (i.e., communication), throughout the HTx process and the care afterward. The healthcare team played a great role in patients’ compliance with the immunosuppressive treatment and their mental health. *“P9: they give me indications, instructions, advice, directives that I apply to the letter.”*

However, the burden and the side effects of immunosuppressive treatment had a negative effect firstly on compliance as well as mental health. Most recipients accepted this as a necessity because they linked it to the graft survival. “*P8: It’s part of my life”. “P9: It’s these immunosuppressants that keep me alive. Because of my graft*.”

To counterbalance these negative effects, recipients called on internal resources such as strength of character and coping skills (i.e., faith, hope, goals, optimism, positive thinking, humor). This resource helped them face difficulties such as anxiety and depression, fears and concerns about the future, survival and graft rejection, and side effects of immunosuppressive treatment. Although, calling on internal resources had a direct and positive impact on the HRQoL, some patients did not have this ability. *“P14: It’s true that [Transplantation] was hard physically, physically and more than morally, because morally I always held on”*.

Return to work could positively affect the HRQoL by enhancing the social role and physical and mental activity. However, some financial aspects could disturb the HRQoL. *“P3: I have the chance to work because it’s an opportunity, right? It is a way to resume a normal social life and be reintegrated into society”*.

Heart recipients had mixed feelings about their donor and donation process. For some, it could affect their mental health by causing guilty. *“P6: Regarding the donor, I have a lot of gratitude towards the donor, but I am not at all trying to get into the drama… because there was necessarily a drama”*.

## DISCUSSION

The quantitative approach aims to identify relationships between variables, to explain or predict phenomena, with the goal of generalizing the results to a broader population. In contrast, the qualitative approach focuses on understanding and interpreting meanings specific to the context in which the data are collected (22).

This qualitative study investigated how HTx recipients perceived their HRQoL and factors that affected it. We used an inductive qualitative method to identify areas of HRQoL and its determinants that HTx recipients considered important. This qualitative study provides new insights into the experiences and perspectives that define HTx recipients’ HRQoL. Identifying these different themes could help recommending a relevant measurement questionnaire to be used in HTx registry.

As in others studies, most patients reported positive experiences with HTx (23,24). The improvement in HRQoL was due to factors such as family support, the return to a certain physical autonomy, the absence of disabling side effects and finally the return to a quasi-normal social life.

As in other studies, most patients reported that the mental health dimension was important throughout the HTx experience, both pre and post-transplant (25). During these periods, emotional tensions appeared such as fear of dying, then the joy of the call for the HTx, then the anxiety caused by the side effects and the fear of rejection. As in other studies, depression and anxiety affected not only patients waiting for HTx but also HTx recipients (26). Healthcare professionals should consider these symptoms because they are associated with complications such as increased incidence of stroke and septicemia after HTx (26). The most commonly used questionnaires for assessing anxiety and depression are the Hospital Anxiety and Depression Scale (HADS) and the Beck Depression Inventory (BDI) (27,28). They are available in French and are widely used in heart transplant populations (29,30).

Most recipients declared a good physical condition and physical capacity. Depending on their physical condition, they were able to resume many of their previous daily life activities. However, the whole group reported different levels of fatigue and pain (31). Fatigue was previously reported to be common and distressing after HTx and thus may affect recipients’ ability to return to work or a social activity and be a barrier to daily life (31). Unlike the pre-transplant period featuring shortness of breath (dyspnea), this symptom was not mentioned during interviews with recipients (32).

As a clinical determinant, the severity of heart disease has a strong impact on HRQoL before transplantation. The transplantation does not solve everything. Some patients, particularly those with extracardiac involvement, may continue to have symptoms even after the transplantation. Patients who were more severely ill before HTx were less satisfied with their lives and felt they were not doing as well; they experienced more family-related stress and used more negative coping strategies than less severely ill patients (10).

The Minnesota Living with Heart Failure Questionnaire (MLHFQ) and the Kansas City Cardiomyopathy Questionnaire (KCCQ) are two validated tools used to assess QoL in patients with cardiomyopathy, including HTx recipients (33,34) . They evaluate physical limitations such as dyspnea, fatigue, edema, chest pain and activity restrictions. Both questionnaires have been validated in French and are widely used in the context of HTx (35). Given their comprehensive assessment of symptoms and functional limitations, these questionnaires would be appropriate for evaluating QoL in HTx recipients.

This study provided an understanding of the link recipients perceived concerning the immunosuppressive treatment, the risk of graft rejection and their HRQoL. Having become very effective, immunosuppressive treatment nevertheless presents serious disadvantages such as side effects, sensitivity to opportunistic infections or tumors. Patients may become discouraged by the burden of side effects, causing them to feel worried, anxious and uncertain about the impact on their body (36). Although rejection was a major concern for recipients, side effects influenced compliance with the immunosuppressive regimen (37). Issues regarding medication administration and medication-associated adverse effects emerged as major barriers to good HRQoL. Adherence to the immunosuppressive regimen could be enhanced by a good healthcare team relationship (23) and therapeutic education (38). Recipients can become dissatisfied with the HTx because of lack of necessary information about transplantation, particularly the side effects of treatment.

The Medication Experience Scale for Immunosuppressants (MESI) and the Modified Transplant Symptom Occurrence and Symptom Distress Scale (MTSOSD-59R) are two questionnaires validated in French for HTx recipients. The MESI assesses adherence and recipients experiences with immunosuppressive medications, while the MTSOSD-59R evaluates the occurrence and the distress associated with side effects from these treatments. Both questionnaires could be utilized within a HTx registry (13,39,40).

Our results are consistent with previous works also finding very strong relationships between the transplant recipient and the healthcare team (41). Recipients considered following the healthcare team’s recommendations as a necessary condition for everything to go well.

As shown previously, the relationship between the recipient and the healthcare team is one of unbalanced dependence (23). Poor relationships can result in reduced compliance with the postoperative regimen (41) and a strong commitment in the relationship with the healthcare team leaves patients dependent on their HTx and follow-up teams. Nevertheless, this relationship seems widely accepted and assumed by the patients.

One essential outcome of this study was the importance of external support. Heart recipients resorted to social support, calling upon family or healthcare professionals as an external resource to assist in addressing a problem or in helping manage a stressful situation. All recipients emphasized the importance of family particularly during the most difficult moments of the HTx. Therefore, family support is a strong determinant of HRQoL. In some cases, relationships were strengthened, and in others, relationships weakened. Some studies have shown a burden of care and exhaustion among caregivers (spouse, children) (42).

Internal resources helped recipients overcome HTx obstacles, reduce stress and improve perceived their HRQoL. Recipients used several styles of internal resources or coping strategies to face the difficulties encountered. For example, being humorous, such as laughing at themselves and joking to play down a critic situation, was viewed as one type of coping mechanism that can have positive effects and reduce the burden of perceived stress (43). Several recipients mentioned a sens of humor as necessary to cope with the difficulties of life with a transplant. Another example is acceptance or resignation previously found effective in reducing perceived stress in the presence of stressful events (44).

Many recipients searched for additional information about their condition, the drugs and their side-effects. This search seemed to help them cope with the situation and live with their condition (45). For example, recipients who knew about their medications were in a better position to ask questions and were able to solve problems and discuss strategies to cope with side effects (46). Certain coping strategies can influence adherence to medical treatment (47).

Despite the difficulties encountered, recipients were determined to remain positive and generally coped well using some combination of coping strategies when dealing with stressful situations (45). Taking into account the ability to mobilize internal resources could help physicians determine a patient’s needs for additional support, such as referral to a psychologist.

To assess the coping strategies employed by HTx recipients, the Brief COPE, translated in French, a widely used instrument for identifying individual coping strategies, could be utilized (48–50).

Aspects related to the donor or socio-economic considerations did not play a big part in recipients’ HRQoL because they did not spontaneously address these subjects. Aspects related to the graft and the donor can be assessed using the Transplanted Organ Questionnaire (TOQ), which explores the recipient’s experience with their transplanted organ (51).

Several points in the present study must be considered when drawing conclusions about our findings. Similar to any qualitative study, our results cannot be generalized to another population. The interviews were conducted in French and then translated to English for the present paper; therefore, loss of meaning during translation cannot be ruled out. We assumed that patients would not spontaneously discuss certain intimate topics. Thus, because our interview guide did not provide them in advance, they might not be addressed in our study. For example, recipients did not spontaneously mention problems related to sexual dysfunction. However, multiple studies have shown a higher prevalence of sexual dysfunction in men and women compared with the general population (52,53).

## Conclusion

In this study, we identified several key areas that contribute to the HRQoL of HTx recipients. Mental health, physical capacity, symptoms and comorbidities, social support, coping strategies and autonomy seemed important and should be considered in the selection of tools for assessing HRQoL.

While existing PROMs do not comprehensively assess all domains of physical, psychological, and social QoL following HTx, a sequential assessment strategy could be recommended. This approach would combine a generic QoL instrument, , with condition- or HTx specific tools , tailored to specific research objectives. This strategy, although more complex to implement, allows to capture the broad and specific aspects of the HTx recipients’ experience.

## Data Availability

All the data used and referenced in this manuscript are available and can be requested from the authors

## Acknowledgement

We would like to extend our warmest thanks to the patients and healthcare professionals who agreed to take part in this study at the following facilities: Henri Mondor (Paris), Pitié Salpetrière (Paris), Laennec (Nantes), Arnaud De Villeneuve (Montpellier), Louis Pradel HCL (Lyon).

## Authorship

Redouane Mahmoudi: Participated in the conception of the work, in the interviews, data analysis, in interpretation of data and in the writing of the paper.

Pascal Battistella: Participated in interpretation of data and in the writing of the paper. Laurent Sebbag: Participated in interpretation of data and in the writing of the paper. Latame Komla Adoli: Participated in interpretation of data and in the writing of the paper.

Francis Guillemin: Participated in the conception of the work, in interpretation of data and in the writing of the paper.

Cécile Couchoud: Participated in the conception of the work, data analysis, in interpretation of data and in the writing of the paper.

All authors approved the final version of the paper.

